# Reappearance of Effector T Cells Predicts Successful Recovery from COVID-19

**DOI:** 10.1101/2020.05.11.20096263

**Authors:** Ivan Odak, Joana Barros-Martins, Berislav Bošnjak, Klaus Stahl, Sascha David, Olaf Wiesner, Markus Busch, Marius M Hoeper, Isabell Pink, Tobias Welte, Markus Cornberg, Matthias Stoll, Lilia Goudeva, Rainer Blasczyk, Arnold Ganser, Immo Prinz, Reinhold Förster, Christian Koenecke, Christian R Schultze-Florey

## Abstract

**Background:** Elucidating the role of T cell responses in COVID-19 is of utmost importance to understand the clearance of SARS-CoV-2 infection.

**Methods:** 30 hospitalized COVID-19 patients and 60 age- and gender-matched healthy controls (HC) participated in this study. We used two comprehensive 11-color flow cytometric panels conforming to Good Laboratory Practice and approved for clinical diagnostics.

**Findings:** Absolute numbers of lymphocyte subsets were differentially decreased in COVID-19 patients according to clinical severity. In severe disease (SD) patients, all lymphocyte subsets were reduced, whilst in mild disease (MD) NK, NKT and γδ T cells were at the level of HC. Additionally, we provide evidence of T cell activation in MD but not SD, when compared to HC. Follow up samples revealed a marked increase in effector T cells and memory subsets in convalescing but not in non-convalescing patients.

**Interpretation:** Our data suggest that activation and expansion of innate and adaptive lymphocytes play a major role in COVID-19. Additionally, recovery is associated with formation of T cell memory as suggested by the missing formation of effector and central memory T cells in SD but not in MD. Understanding T cell-responses in the context of clinical severity might serve as foundation to overcome the lack of effective anti-viral immune response in severely affected COVID-19 patients and can offer prognostic value as biomarker for disease outcome and control.

**Funding:** Funded by German Research Foundation, Excellence Strategy – EXC 2155 “RESIST”–Project ID39087428, and DFG-SFB900/3–Project ID158989968, grants SFB900-B3 to R.F., SFB900-B8 to I P. and C.K.

## Introduction

Cross-species infections are becoming more common due to increase in interlinking human and animal activities, furthered by great genetic diversity of viruses^1^. The threat of emerging global pandemics by new virus species and/or mutants has been significantly heightened in the wake of severe acute respiratory syndrome coronavirus (SARS-CoV)^2^ and Middle East respiratory syndrome coronavirus (MERS-CoV)^3^ outbreaks. Both of these zoonotic coronaviruses are highly infectious and have high fatality rates^4^. In December 2019, another type of coronavirus, later named SARS-CoV-2, was reported in patients with pneumonia, causing coronavirus disease 2019 (COVID-19)^5^. SARS-CoV-2 has since turned into a global pandemic with a total of 3 925 815 confirmed cases of COVID-19 and 274 488 fatal cases as of May 10^th^ 2020 according to recent WHO situation report^6^.

Since the onset of the SARS-CoV-2 pandemic, research focuses on immune profiling of the infection, striving towards better patient management and treatment. As it is the case in the majority of viral infections, key players in battling the SARS-CoV-2 appear to be NK, T and B cells^7,8^. According to available data^9–11^, COVID-19 patients with severe disease have increased levels of C-reactive protein and IL-6 as well as augmented neutrophil counts but reduced lymphocyte counts in blood. Moreover, NK cells and CD8^+^ T cells in COVID-19 patients appear to be functionally exhausted, indicated by increased expression of NKG2A and lower production of IFN-γ, TNF-α and IL-2^12^. Nevertheless, it is unclear whether and how profiling of T cell responses can be used as prognostic biomarker for disease outcome and control. Furthermore, no data is available on the role of γδ T cells in anti-SARS-CoV-2 immune responses, although it has been demonstrated that these cells contribute to immunity against SARS-CoV and other viruses^13–15^.

In the present study we analysed dynamics of NK, NKT, αβ and γδ T cells subsets in the peripheral blood of patients with mild and severe COVID-19 compared to gender- and age-matched controls. To reliably assess major lymphocyte subsets’ profiles during successful immune response against SARS-CoV-2 infection, we developed two comprehensive Good Laboratory Practice (GLP)-conforming 11-color flow cytometric panels approved for clinical diagnostics. Using those panels, we examined the composition of seven major lymphocyte populations in patients with mild and severe COVID-19 and followed formation of effector and memory αβ and γδ T cells from consecutive blood samples of patients who did or did not clinically improve. We found that recovery from COVID-19 was closely associated with expansion and differentiation/maturation in αβ, but not γδ T cells.

## Materials and Methods

### Study participants

Patients with PCR-confirmed SARS-CoV-2 infection were recruited at Hannover Medical School from March 30th until April 16th 2020. Based on the clinical presentation, disease was classified as “mild” or “severe” for every single patient at admission. Mild disease was defined for patients with stable lung parameters with no oxygen flow or of up to 3 liters per minute. In contrast, severe disease was defined as oxygen flow equal or greater than 6 liters per minute to maintain a SpO_2_ > 90%, or non-invasive or invasive ventilation. Patient characteristics are shown in **Table 1**. To assess the impact of infection on lymphocyte subsets, age- and gender-matched healthy controls (HC) were selected for every patient in a 2:1 control-to-patient ratio. Those patients of 56 years of age and older were gender-matched to the group of 56-69 year old healthy controls. Healthy controls were recruited through the Institute of Transfusion Medicine in October and November 2019, prior to SARS-CoV-2 outbreak. Healthy control characteristics are listed in **Supplementary Table 1**. The study was approved by the institutional review board at Hannover Medical School (#9001_BO_K2020 and #8606_BO_K2019) and informed consent was obtained from all patients and healthy controls.

**Table 1:** Patients’ Characteristics. Abbreviations: PEMC: pre-existing medical conditions, cv: cardio vascular, m: male, f: female, NA: not available, *: days after onset of symptoms

| ID | Severity | Age | Gender | PEMC | cv PEMC | Symptoms | First Sampling* | Con-<br>valescence | Convalescence<br>Sampling* |
| --- | --- | --- | --- | --- | --- | --- | --- | --- | --- |
| 001 | mild | 71 | m | no | no | Fever, malaise | 13 | NA | NA |
| 002 | mild | 68 | m | yes | yes | Fever, diarrhea | 13 | NA | NA |
| 003 | mild | 64 | m | yes | yes | Malaise | 14 | NA | NA |
| 012 | mild | 48 | f | yes | no | Fever, dry cough, dyspnea | 8 | NA | NA |
| 013 | mild | 85 | m | yes | yes | Fever, dry cough | 4 | yes | 17 |
| 014 | mild | 51 | m | yes | yes | Fever, myalgia | 16 | NA | NA |
| 016 | mild | 91 | m | yes | yes | Dry cough, malaise | 9 | no | 12 |
| 019 | mild | 48 | f | yes | no | Dry cough | 9 | NA | NA |
| 020 | mild | 57 | f | yes | yes | Dyspnea, myalgia, diarrhea, emesis | 8 | NA | NA |
| 021 | mild | 48 | m | yes | yes | Fever, dry cough, myalgia | 3 | NA | NA |
| 022 | mild | 75 | m | yes | yes | Fever | 8 | no | 18 |
| 023 | mild | 60 | m | yes | yes | Dry cough, myalgia, cephalgia | 10 | NA | NA |
| 027 | mild | 83 | f | yes | yes | Fever, dry cough, dyspnea, malaise | 5 | yes | 12 |
| 028 | mild | 42 | m | yes | no | Nausea, emesis | 3 | NA | NA |
| 031 | mild | 79 | m | yes | yes | Diarrhea, nausea, emesis | 2 | no | 7 |
| 004 | severe | 75 | f | yes | yes | Fever, dry cough, dyspnea | 7 | yes | 17 |
| 005 | severe | 60 | m | yes | yes | Fever, dry cough | 20 | NA | NA |
| 006 | severe | 51 | m | yes | no | Fever, diarrhea, nausea | 12 | yes | 19 |
| 007 | severe | 60 | m | yes | no | Fever, dry cough, diarrhea | 16 | no | 33 |
| 008 | severe | 56 | m | yes | yes | Fever, dry cough, dyspnea | 19 | no | 26 |
| 009 | severe | 68 | m | yes | no | Dry cough, diarrhea | 18 | yes | 25 |
| 010 | severe | 69 | m | yes | yes | Dry cough, dyspnea | 21 | no | 38 |
| 011 | severe | 45 | m | yes | yes | Dry cough, dyspnea | 5 | no | 12 |
| 015 | severe | 47 | m | yes | yes | Fever, dry cough, dyspnea | 16 | no | 31 |
| 017 | severe | 19 | m | yes | no | Malaise | 5 | yes | 19 |
| 018 | severe | 82 | f | yes | yes | Dry cough | 7 | NA | NA |
| 025 | severe | 65 | m | yes | yes | Fever, dry cough, dyspnea | 8 | yes | 16 |
| 026 | severe | 71 | m | yes | yes | Fever, dry cough | 13 | no | 18 |
| 029 | severe | 53 | m | yes | no | Fever, dry cough, diarrhea, nausea | 8 | no | 15 |
| 030 | severe | 39 | m | no | no | Fever, dyspnea | 32 | NA | NA |

### Flow cytometry

EDTA-anticoagulated whole blood was stained with the antibodies as listed in **Supplementary Table 2**. For analysis of absolute cell counts BD TruCount™ tubes were used, and cells were analyzed in a lyse-no-wash fashion according to manufacturer’s instructions. For analysis of activation markers, cells were analyzed in a lyse-wash fashion. Stained cells were acquired on BD FACS Lyric, standardized for clinical cell analysis. Flow cytometry data were analysed using FCSExpress 7 (De Novo Software).

### Statistical analysis

Data were analyzed using Prism 7.05 (GraphPad). For group comparisons two-tailed student t test or Mann-Whitney U test were used where applicable; for longitudinal changes two-tailed paired t test was used.

## Results

Our cohort of 30 hospitalized COVID-19 patients was predominantly male (80%), of older age (mean 61 years, range 19-91) and 93% had mostly mild pre-existing medical conditions. Patients were classified upon admission as mild (n=15) and as severe (n=15, **Table 1**). Patients with severe disease (SD) were slightly older and had a higher proportion of male gender, although both not significantly different from patients with mild disease (MD). Characteristics of healthy controls (HC) are listed in **Supplementary Table 1**.

### Severe COVID-19 patients show reduced counts of lymphocyte subsets

Using BD TruCount™ technology, we observed significantly reduced lymphocyte numbers in COVID-19 patients, inversely correlated with disease severity (**Supplementary Fig. 1**). To further characterize the composition of the lymphocyte pool, we developed two comprehensive GLP-conforming 11-color comprehensive flow cytometric mAb panels, which were approved in-house for clinical diagnostics. The first panel of mAb used in this study (Supplementary Fig. 2A) allowed us to profile NK, NKT, B, αβ and γδ T cell numbers in blood of HC and COVID-19 patients with MD or SD. Compared to MD patients, we found strikingly lower numbers of NK, NKT, γδ-, and CD8^+^ cells in the blood of SD COVID-19 patients (**Fig. 1**). γδ^+^Vγ9^+^ cells are reported to be responsive to phospho-antigens^16,17^ and we noticed a trend to reduced numbers of γδ^+^Vγ9^+^ in SD patients compared to MD (P=0.0502, two-tailed Student’s t test). In addition, all subsets investigated were significantly reduced in SD patients compared to HC (**Fig. 1**). Interestingly, MD patients had similar levels of NK, NKT, γδ-, conventional CD4^+^ (CD4_conv_) and CD8^+^ cells as compared to HC. Of note, both MD and SD patients showed a marked reduction of regulatory T cells (**Fig. 1**). Together, these data provide a detailed depiction of reduced lymphocyte subsets in mild and severe COVID-19 patients, employing certified counting beads for analysis of absolute cell numbers.

**Figure 1:**
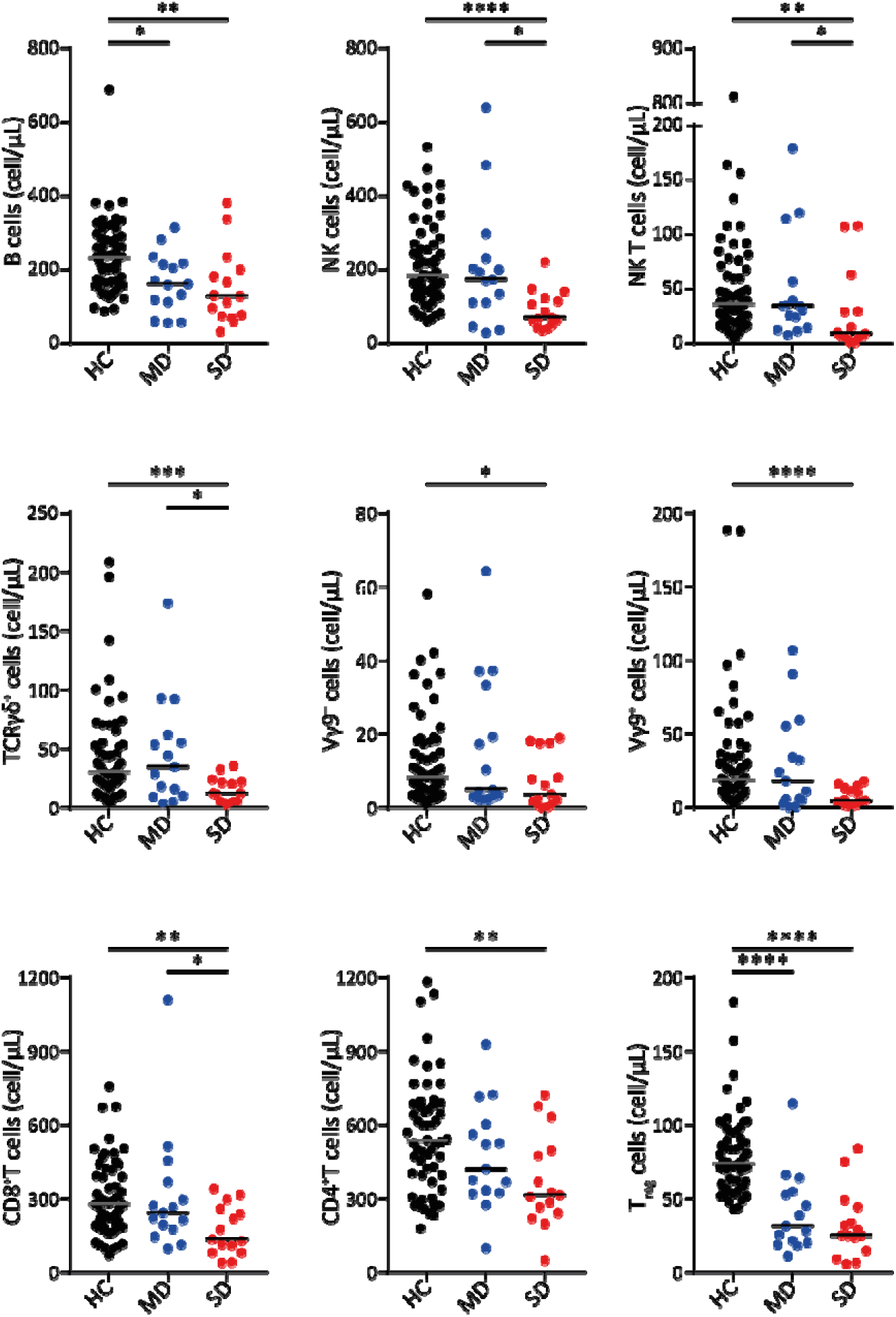
Absolute numbers of lymphocyte subsets in healthy controls and mild and severe COVID-19 patients. Lines represent median. Statistical analysis was performed by Mann-Whitney U-test or Student’s t test where applicable. *p< 0.05, **p < 0.01, ***p < 0.001, ****p < 0.0001; ns: not significant; HC: Healthy Control; MD: Mild Disease; SD: Severe Disease.

### Patients with severe COVID19 infection lack generation of effector and central memory αβ CD4_conv_ and CD8^+^ cells

To characterize the involvement of different subsets of αβ CD4_conv_, αβ CD8^+^, and γδ T cells based on their antigen experience^18,19^, we developed a staining panel dedicated to identifying four distinct populations based on CD62L and CD45RA expression (**Supplementary Fig. 2B**). Looking at the distribution of CD45RA^+^CD62L^+^ on conventional CD4^+^ T cells (CD4_conv_) we defined naïve (CD4_naïve_, CD45RA^+^CD62L^+^), effector/effector memory (CD4_eff/em_, CD45RA^-^CD62L^-^), terminally differentiated cells (CD4_temra_, CD45RA^+^CD62L^-^) and central memory (CD4_cm_, CD45RA^-^CD62L^+^). Based on this allocation, we observed a marked decrease of CD4_eff/em_ in SD compared to MD patients (**Fig. 2A**). In addition, both COVID-19 groups had increased frequencies of CD4_temra_ and decreased CD4_eff/em_ compared to HC (**Fig. 2A**). The same observations were made when absolute cell counts were calculated (**Supplementary Fig. 3A**). Investigating the memory phenotypes of αβ CD8^+^ cells revealed a striking increase in frequencies of CD8_eff/em_ cells, as well as a decrease of CD8_naïve_ T cells in MD compared to HC and SD (**Fig. 2B**). These observations indicate successful mobilization and/or differentiation of cytotoxic lymphocytes (CTL) in MD patients. Compared to MD, patients with SD had higher frequency of CD8_naïve_ T cells and lower frequency of CD8_eff/em_ cells, implying deficits in successfully mounting CTL responses in SD patients or increased recruitment to the peripheral organs. Furthermore, we found similar frequencies of CD8_cm_ and CD8_temra_ in samples from MD and HC but a decrease of these cell populations in SD (**Fig. 2B**), again hinting towards an impaired CTL response in patients with SD. The absolute numbers of CD8 memory subsets was changed similarly with the exception of CD8_naïve_ T cells in SD which were similar to those of MD patients (**Supplementary Fig. 3B**).

**Figure 2:**
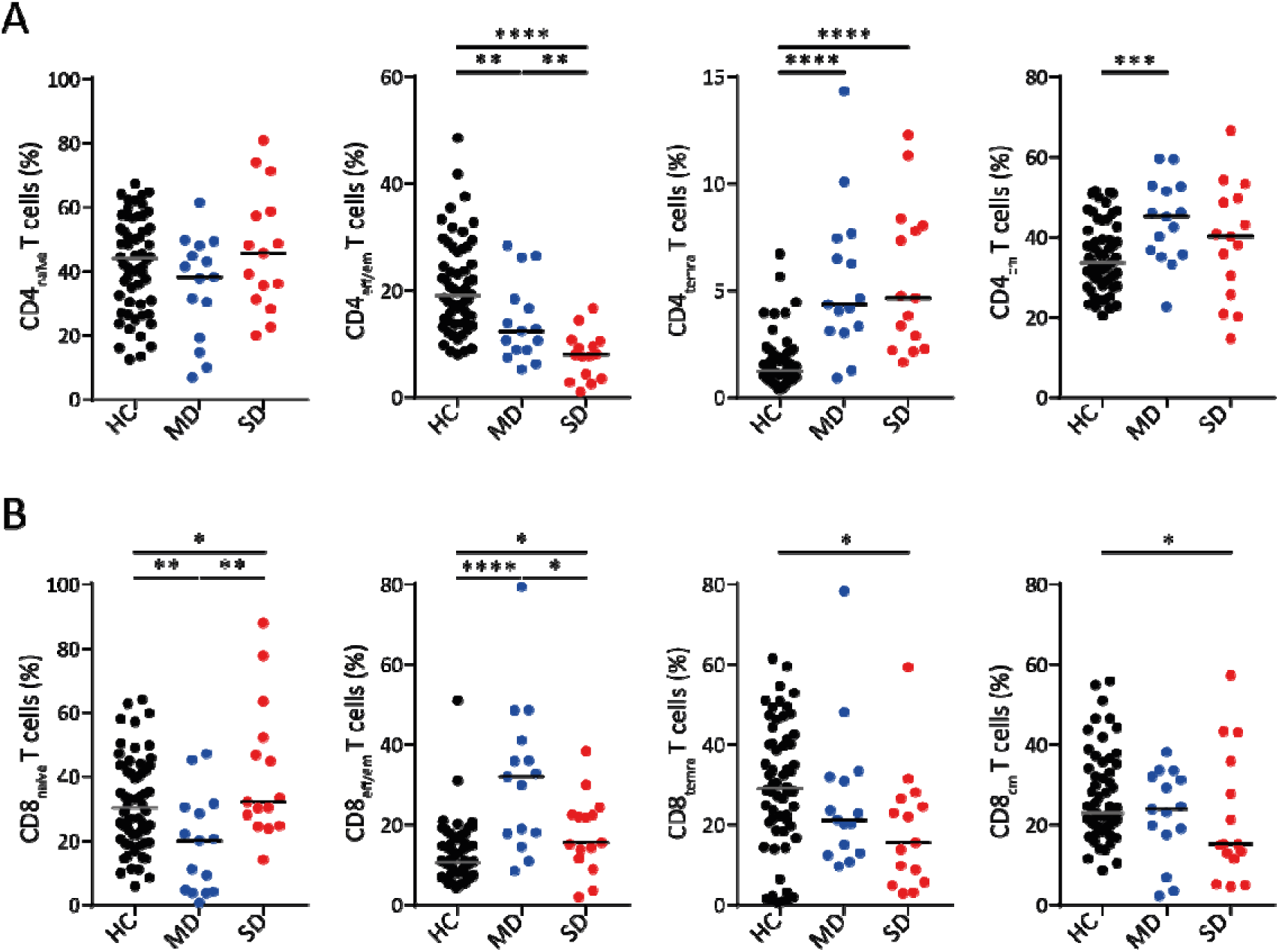
Mild COVID-19 disease is paralleled by increased levels of effector and memory T cells. (A) Gated on CD4_conv_. (B) Gated on CD8^+^ T cells. Lines represent median. Statistical analysis was performed by Mann-Whitney U-test or Student’s t test where applicable. *p< 0.05, **p < 0.01, ***p < 0.001, ****p < 0.0001; ns: not significant; HC: Healthy Control. MD: Mild Disease; SD: Severe Disease.

### γδ T cells from COVID-19 patients exhibit a distinct memory signature

Human γδ T cells have been shown to be important for regulating immune responses to CMV, HCV and SARS-CoV infections^13–15^. Using a panel of 11 specific mAb (**Supplementary Fig. 2B**), we found that expression of CD45RA and CD62L on γδ cells would also allow their categorization into four subsets as defined above for αβ T cells. Interestingly, we revealed a marked increase in both frequencies and absolute cell numbers of naïve-like γδ (γδ_naïve–l_) cells, and a proportional decrease in effector-like γδ (γδ_eff-l_) cells in both groups of patients suffering from COVID-19 (**Fig. 3**). These data suggest that γδ_eff-l_ T cells might be recruited from the blood to peripheral organs such as the lungs of COVID-19 patients and that these cells might participate in the immune response against SARS-CoV-2 infection.

**Figure 3:**
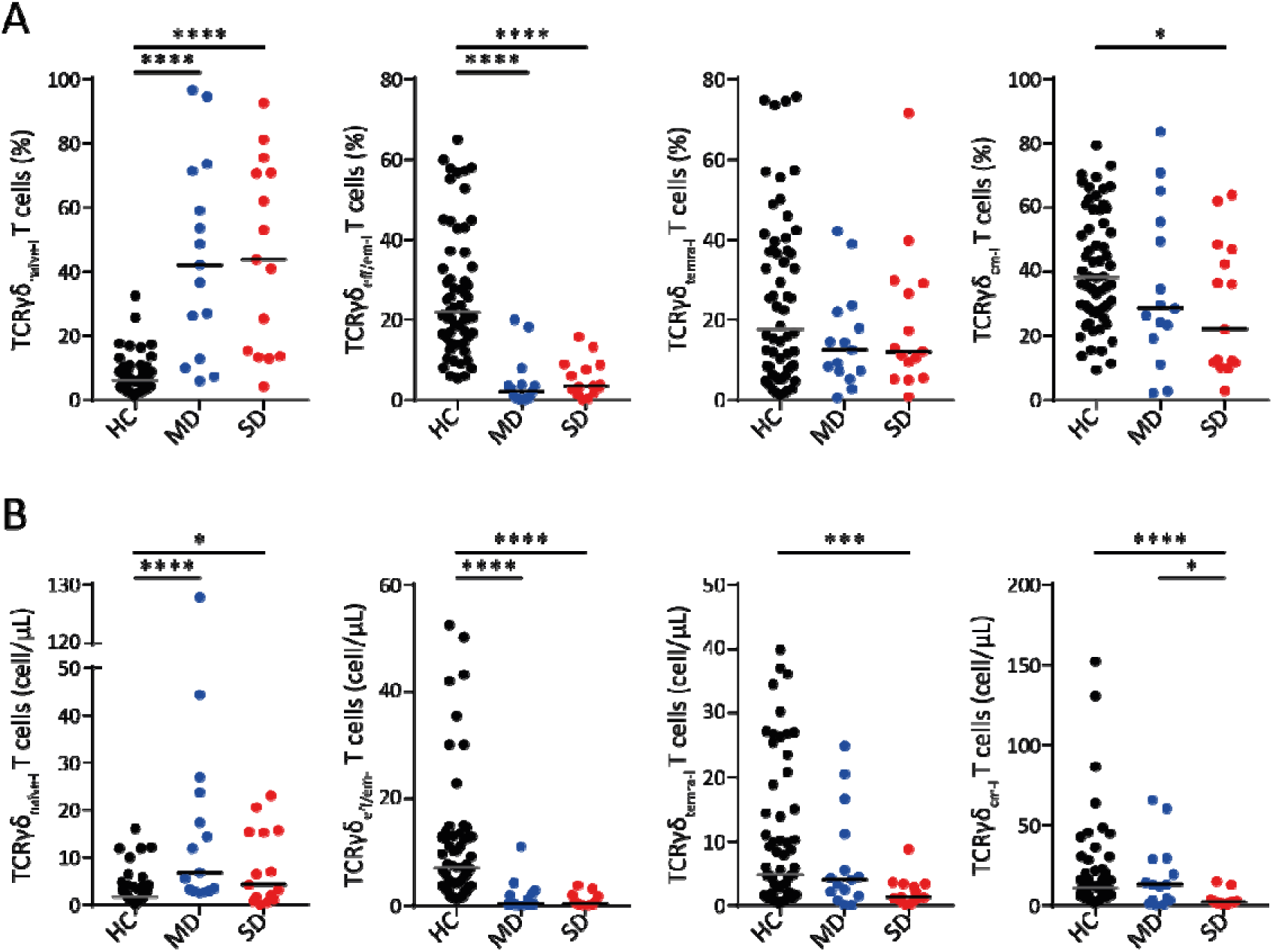
γδ T cells from COVID-19 patients exhibit a distinct memory signature. (A) Gated on γδ T cells. (B) Absolute numbers of γδ T cell subpopulations. Lines represent median. Statistical analysis was performed by Mann-Whitney U-test or Student’s t test where applicable. *p< 0.05, **p < 0.01, ***p < 0.001, ****p < 0.0001; HC: Healthy Control. MD: Mild Disease; SD: Severe Disease.

### Early establishment of T cell memory suggests better clinical outcome

To evaluate the hypothesis that convalescence from COVID-19 is related to establishment of successful T cell immunity, we evaluated the signature of CD4_conv_ and CD8^+^ cells based on the clinical course of the disease. To that end, we collected longitudinal samples of 5 MD and 12 SD patients. We defined convalescence as an improvement of lung function (e.g. reduction of oxygen flow or ventilation parameters), excluding transient variations, thus only taking long-lasting clinical improvement in consideration. Convalescence rate in MD and SD patients occurred at similar frequencies (2/5 vs 5/12) while the period of follow-up was slightly longer in SD patients without reaching statistical significance (SD: 10.08 vs MD: 7.60 days; **Table 1**). Therefore, we re-grouped the MD and SD patients into convalescing (n=7) and non-convalescing patients (n=10). In all convalescing patients irrespective of the disease severity, we observed a strong decrease of the frequency of CD4_naïve_ and a marked increase of CD4_eff/em_ and CD4_cm_ populations (**Fig. 4A**) suggesting a turnover of naïve to memory/effector cells. The same observations were made for all convalescing patients regarding the relative distribution of naïve and memory CD8^+^ T cell subsets (**Fig. 4B**). Notably, no significant changes could be observed for any of the CD4_conv_ and CD8^+^ subpopulations in non-convalescing patients (**Fig. 4**). We also failed to observe any significant alterations in frequencies of the different subtypes of γδ cells in relation to disease clearance (**Supplementary Fig. 4**). We therefore suggest that the alterations observed for CD4_conv_ and CD8^+^ αβ T cells in convalescing patients that recover from COVID-19 are linked to differentiation of effector/memory αβ T cells.

**Figure 4:**
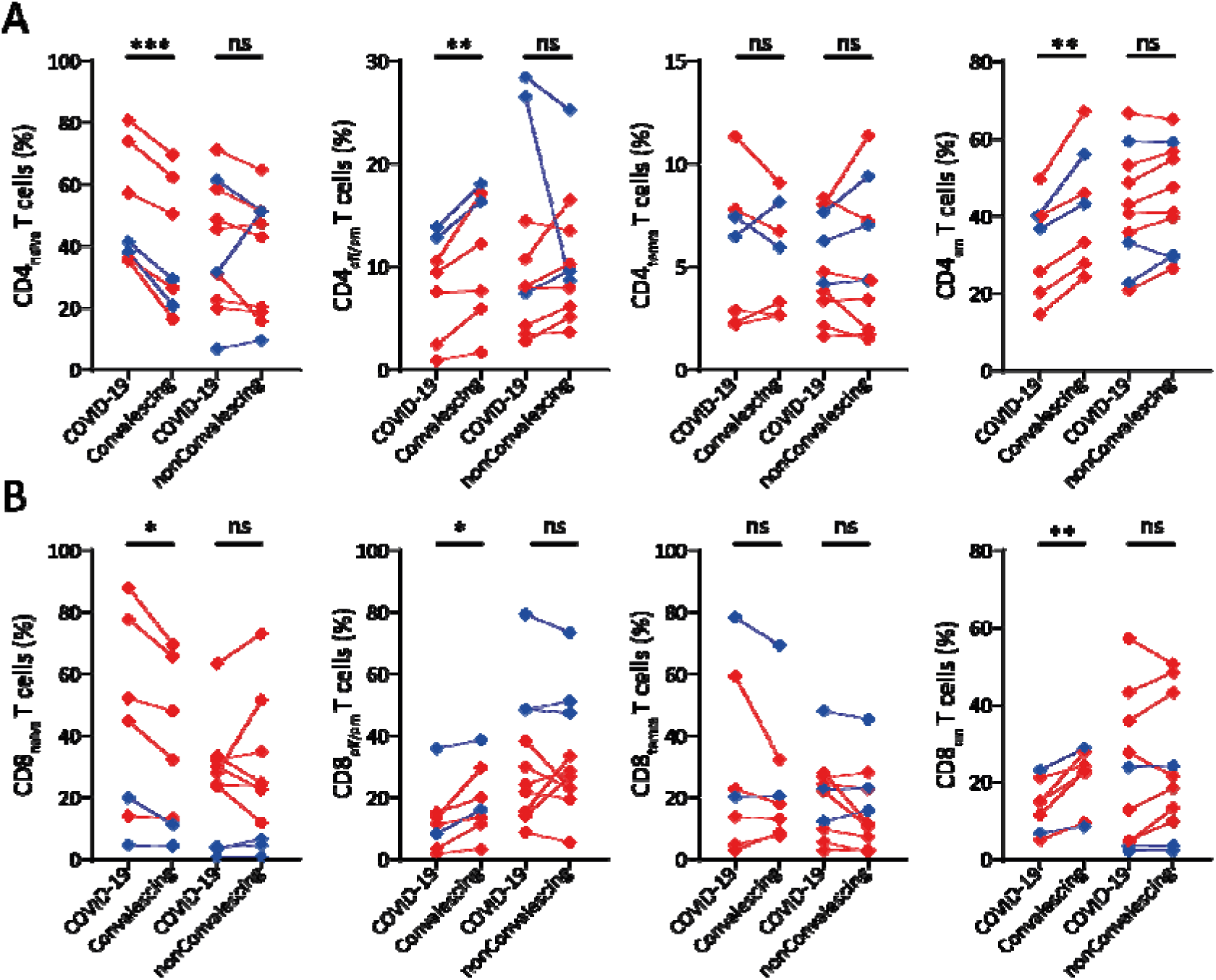
Increase of effector/memory T cells suggests better clinical outcome. (A) Gated on CD4_conv_. (B) Gated on CD8^+^. Blue diamonds represent mild disease and red diamonds severe disease patients upon admission. Statistical analysis was performed paired Student’s t test. *p< 0.05, **p < 0.01, ***p < 0.001; ns: not significant.

## Discussion

From an immunological perspective, adaptive anti-viral responses occur in two layers. First, a NK, NKT and CD8^+^ T cell response is programmed to prevent the disease to proceed to a severe phase. Second, helper T cells are poised to influence and program B cells for specific neutralizing antibody formation, conferring long-lasting humoral immunity^20^. The establishment of immunity to SARS-CoV-2 has been associated with the presence of neutralizing IgG antibodies in blood sera^21,22^. However, some viruses such as influenza are notoriously known to be resilient to establish humoral immunity^23^. This might also be the case for SARS-CoV-2. We therefore studied changes in T cell subsets in COVID-19 patients.

Applying two comprehensive GLP-conforming 11-color flow cytometric panels approved for clinical diagnostics, this prospective single-center study from Germany identified profound changes in absolute numbers in seven different lymphocyte subsets in hospitalized COVID-19 patients with mild and severe disease. These findings were in line with and extended the current literature on COVID-19-induced lymphocytopenia^9–11^. Additionally, we revealed changes in the formation of different T memory cell subsets comparing mildly and severely affected COVID-19 patients with healthy controls. Moreover, in a longitudinal approach we showed that convalescing – but not non-convalescing – patients develop increased frequencies of different subsets of CD4_conv_ and CD8^+^T cell showing memory/effector phenotypes. Together, our findings indicate that mounting an efficient T cell response allows hospitalized patients to combat and overcome SARS-CoV-2 infections.

In agreement with recent reports, we observed a general decrease of lymphocytes in all COVID-19 patients that was more pronounced in severe cases^8–10^. Such a reduction of immune cells in the blood of SD patients might be due to several causes. It is conceivable that infection with SARS-CoV-2 directly affects differentiation and/or production in and release from primary lymphoid organs. Although this virus in general is not suspected to infect and destroy immune cells or their progenitors to a large degree, it seems possible that such effects might be secondary and caused by massive release in inflammatory mediators as a response to the infection^9,10^. Alternatively, failure to observe lymphocytes in the blood might be due to their homing/recruitment to the lung or other infected organs^11^. COVID-19 patients with a mild clinical course did not show reduction in NK, NKT and γδ T cells counts. NK cells are part of the innate immunity and their significance in clearance of viral infections has been well documented^24,25^. It is thus reasonable to assume that these cells contribute to keeping the disease from progressing, as suggested before^8^. The observation that these innate and innate-like immune cells are present in regular numbers at early stages of the disease might also indicate that this arm of the immune system helps to contain early virus infection.

Compared to HC, COVID-19 patients showed already altered effector/effector memory and naïve T cell proportions at hospital admission. Increased frequencies of effector and memory populations in MD, but not SD, suggest that MD patients already launched a broad adaptive immune response at that time point, whilst this was presumably not the case in severely affected patients. Along the same line, a shift towards further activation of αβ T cells coincided with convalescence, also hinting at the presence of SARS-CoV-2 responsive T cells. These findings are in line with very recent reports describing the role of T cell immunity in COVID-19 patients^26–29^.

Data of the present study highlight the role of T cells in successfully controlling SARS-CoV-2 infections and holds implications for vaccine design and assessment of successful immune response in vaccine trials. Furthermore, comprehensive lymphocyte subset analyses by flow cytometry can be easily implemented and might serve as a biomarker for disease outcome and control.

## Data Availability

Data are made available upon written request to the corresponding author after signing a data transfer agreement.

## Acknowledgements

We thank Jolanta Adolf, Christine Garen, Ellen Hebold, Bianca Krüger and Kerstin Schantl for their support with the experiments. We thank Melanie Drenker and Daniela Garve for their support with the clinical data analysis.

## Author contributions

IO, CK and CRSF designed the study, KS and SD obtained written informed consent from all patients, LG obtained written informed consent from all healthy controls, CRSF and IO performed experiments and analysed experimental data; IO, CRSF, ImP, RF and CK interpreted the data, IO and CRSF wrote the manuscript, JBM, BB, KS, SD, OW, MB, MMH, IsP, TW, MC, MS, LG, RB, AG, ImP, RF and CK reviewed and edited the manuscript.

## References

1 Wong G, Liu W, Liu Y, Zhou B, Bi Y, Gao GF. MERS, SARS, and Ebola: The Role of Super-Spreaders in Infectious Disease. Cell Host Microbe 2015; 18: 398–401.

2 Drosten C, Günther S, Preiser W, et al. Identification of a novel coronavirus in patients with severe acute respiratory syndrome. N Engl J Med 2003; 348: 1967–76.

3 Zaki AM, Van Boheemen S, Bestebroer TM, Osterhaus ADME, Fouchier RAM. Isolation of a novel coronavirus from a man with pneumonia in Saudi Arabia. N Engl J Med 2012; 367: 1814–20.

4 Cui J, Li F, Shi ZL. Origin and evolution of pathogenic coronaviruses. Nat Rev Microbiol 2019; 17: 181–92.

5 Zhu N, Zhang D, Wang W, et al. A novel coronavirus from patients with pneumonia in China, 2019. N Engl J Med 2020; 382: 727–33.

6 WHO. Coronavirus Disease 2019. https://covid19.who.int/ (accessed May 10, 2020).

7 Braun J, Loyal L, Frentsch M, Wendisch D, Georg P. Presence of SARS-CoV-2-reactive T cells in COVID-19 patients and healthy donors. *medRxiv* 2020. DOI:https://doi.org/10.1101/2020.04.17.20061440.

8 Zheng M, Gao Y, Wang G, et al. Functional exhaustion of antiviral lymphocytes in COVID-19 patients. Cell Mol Immunol 2020;: 7-9.

9 Huang C, Wang Y, Li X, et al. Clinical features of patients infected with 2019 novel coronavirus in Wuhan, China. Lancet 2020; 395: 497–506.

10 Yang X, Yu Y, Xu J, et al. Clinical course and outcomes of critically ill patients with SARS-CoV-2 pneumonia in Wuhan, China: a single-centered, retrospective, observational study. Lancet Respir Med 2020; 2600: 1–7.

11 Xu Z, Shi L, Wang Y, et al. Pathological findings of COVID-19 associated with acute respiratory distress syndrome. Lancet Respir Med 2020; 8: 420–2.

12 Qin C, Zhou L, Hu Z, et al. Dysregulation of immune response in patients with COVID-19 in Wuhan, China. Clin Infect Dis 2020; 53: 1689–99.

13 Ravens S, Schultze-Florey C, Raha S, et al. Human γδ T cells are quickly reconstituted after stem-cell transplantation and show adaptive clonal expansion in response to viral infection. Nat Immunol 2017; 18: 393–401.

14 Poccia F, Agrati C, Castilletti C, et al. Anti–Severe Acute Respiratory Syndrome Coronavirus Immune Responses: The Role Played by Vγ9Vδ2 T Cells. J Infect Dis 2006; 193: 1244–9.

15 Ravens S, Hengst J, Schlapphoff V, et al. Human γδ T cell receptor repertoires in peripheral blood remain stable despite clearance of persistent hepatitis C virus infection by direct-acting antiviral drug therapy. Front Immunol 2018; 9. DOI:10.3389/fimmu.2018.00510.

16 Constant P, Davodeau F, Peyrat MA, et al. Stimulation of human γδ T cells by nonpeptidic mycobacterial ligands. Science (80-) 1994; 264: 267–70.

17 Tanaka Y, Tanaka Y, Bloom BR, Morita CT, Brenner MB, Nieves E. Natural and synthetic non-peptide antigens recognized by human γδ T cells. Nature 1995; 375: 155–8.

18 Mangare C, Tischer-Zimmermann S, Riese SB, et al. Robust identification of suitable T-cell subsets for personalized CMV-specific T-cell immunotherapy using CD45RA and CD62L microbeads. Int J Mol Sci 2019; 20. DOI:10.3390/ijms20061415.

19 Martin MD, Badovinac VP. Defining memory CD8 T cell. Front Immunol 2018; 9: 1–10.

20 Shi Y, Wang Y, Shao C, et al. COVID-19 infection: the perspectives on immune responses. Cell Death Differ 2020;: 1451-4.

21 Iwasaki A, Yang Y. The potential danger of suboptimal antibody responses in COVID-19. Nat Rev Immunol 2020;: 1-3.

22 Du L, He Y, Zhou Y, Liu S, Zheng BJ, Jiang S. The spike protein of SARS-CoV - A target for vaccine and therapeutic development. Nat Rev Microbiol 2009; 7: 226–36.

23 Bahadoran A, Lee SH, Wang SM, et al. Immune responses to influenza virus and its correlation to age and inherited factors. Front Microbiol 2016; 7: 1–11.

24 Van Der Vliet HJJ, Pinedo HM, Von Blomberg BME, Van Den Eertwegh AJM, Scheper RJ, Giaccone G. Natural killer T cells. Lancet Oncol 2002; 3: 574.

25 Parham P, Guethlein LA. Genetics of Natural Killer Cells in Human Health, Disease, and Survival. Annu Rev Immunol 2018; 36. D0I:10.1146/annurev-immunol-042617-053149.

26 Weiskopf D, Schmitz KS, Raadsen MP, et al. Phenotype of SARS-CoV-2-specific T-cells in COVID-19 patients with acute respiratory distress syndrome. *medRxiv* 2020;: 2020.04.11.20062349.

27 Ni L, Ye F, Cheng M-L, et al. Detection of SARS-CoV-2-specific humoral and cellular immunity in COVID-19 convalescent individuals. Immunity 2020; published online May. DOI:10.1016/j.immuni.2020.04.023.

28 Grifoni A, Sidney J, Zhang Y, Scheuermann RH, Peters B, Sette A. A Sequence Homology and Bioinformatic Approach Can Predict Candidate Targets for Immune Responses to SARS-CoV-2. Cell Host Microbe 2020; 27: 671–680.e2.

29 Ahmed SF, Quadeer AA, McKay MR. Preliminary identification of potential vaccine targets for the COVID-19 Coronavirus (SARS-CoV-2) Based on SARS-CoV Immunological Studies. Viruses 2020; 12. DOI:10.3390/v12030254.

